# Review of Forecasting Models for Coronavirus (COVID-19) Pandemic in India during Country-wise Lockdowns

**DOI:** 10.1101/2020.08.03.20167254

**Authors:** Abhinav Gola, Ravi Kumar Arya, Animesh, Ravi Dugh

## Abstract

**Background:** COVID-19 is widely spreading across the globe right now. While some countries have flattened the curve, others are struggling to control the spread of the infection. Precise risk prediction modeling is key to accurate prevention and containment of COVID-19 infection, as well as for the preparation of resources needed to deal with the pandemic in different regions.

**Methods:** Given the vast differences in approaches and scenarios used by these models to predict future infection rates, in this study, we compared the accuracy among different models such as regression models, ARIMA model, multilayer perceptron, vector autoregression, susceptible exposed infected recovered (SEIR), susceptible infected recovered (SIR), recurrent neural networks (RNNs), long short term memory networks (LSTM) and exponential growth model in prediction of the total COVID-19 confirmed cases. We did so by comparing the predicted rates of these models with actual rates of COVID-19 in India during the nationwide lockdowns.

**Results:** Few of these models accurately predicted COVID-19 incidence and mortality rates in six weeks, though some provided close results. While advanced warning can help mitigate and prepare for an impending or ongoing epidemic, using poorly fitting models for prediction could lead to substantial adverse outcomes.

**Implications:** As the COVID-19 pandemic continues, accurate risk prediction is key to effective public health interventions. Caution should be taken when choosing different risk prediction models based on specific scenarios and needs. To improve risk prediction of infectious disease such as COVID-19 for policy guidance and recommendations on best practices, both internal (e.g., specific virus characteristics in transmission and mutation) and external factors (e.g., large-scale human behaviors such as school opening, parties, and breaks) should be considered and appropriately weighed.

## 1 Introduction

COVID-19 pandemic, also known as the coronavirus pandemic is currently wreaking havoc in more than 200 countries [1] worldwide. It is caused by Severe Acute Respiratory Syndrome Coronavirus 2 (SARS-CoV-2). As of late July 2020, at least 17 million people are diagnosed with this virus. This infectious disease has left more than 668 thousand people dead. The infection numbers are increasing every day, and without a vaccine, it will be difficult to eradicate this virus.

The virus spreads through contact, mainly through coughing, sneezing, or just being close to an infected person. The droplets released by the coughing or sneezing person can travel several feet and can infect the nearby people. If these droplets fall on nearby surfaces, they can contaminate such surfaces and lead to infecting others coming in contact with such surfaces. Currently, there are no medications to prevent or treat this epidemic [2], but many biotech companies have stepped in the race of developing vaccines with million of dollars at stake.

On January 30, 2020, the first COVID-19 occurrence was registered in India. According to the Ministry of Health and Family Welfare (MoHFW) [3], more than 1.6 million people have been infected by this deadly disease. Coronavirus is also responsible for more than 36,000 deaths until now. India has the highest confirmed case in Asia since June 16, 2020 [1].

Different nations imposed different methods to combat the spread of this disease. These varied for quarantine, mask use, closing of markets/malls, restricting social gatherings, travel restrictions, frequently washing hands, sanitizing frequently used areas, etc.. Some countries imposed lockdowns in different states to restrict the outbreak of the disease further. In India, to counter the spread of this deadly disease, several lockdowns were imposed country-wise. The first nationwide lockdown was imposed on March 24, 2020, for 21 days. The lockdown was further extended on April 14, 2020, till May 3, 2020. Further extensions were imposed on May 3 and May 17, 2020. With a few exceptions, the nation began unlocking on June 1, 2020 [3]. We use the prediction models during these lockdown times. During this time, as the lockdown was at the national level, there were fewer variables in the supposed model. After these lockdowns, India went into partial lockdowns where some states of India were under lockdowns while others were not.

Several prediction models have been used over the past several months to predict the COVID-19 infection rate. These models forecast the rate of infection, recovery, death, or some combination of these three parameters for the COVID-19 patients. The primary evaluation variables can be split into two groups in order to test a forecast model - virus intrinsic: time of incubation, the element of virulence, etc. and outside: size, quarantine, and so on of the infected population. While no model can accurately forecast the rates of infection and mortality, attempts have been made to consider and analyze the strengths and shortcomings of many studies and models presented regarding the coronavirus. Whereas the forecast models used by the health department or Government of India were not disclosed, we can definitely continue with existing models in separate research publications. Each of these models took different approaches and techniques to predict future outbreak rates. In this study, we review major forecasting models that were used in the context of India and compare them with real/true data. In this way, the difference of confirmed cases of COVID-19 between forecasted and true cases gives us the error by which these models made bad predictions. For our study, we take published models before the national unlocking period (i.e. June 1, 2020). All the prediction models were chosen so that they could predict the cases during lockdowns. The future deaths is hard to predict as with a path of hurricane [5]. In the US, the CDC (Centre for Disease Control) is relying on a mash-up of 32 models. Often models are used for projections up to 6 weeks as they rely on a variety of untested assumptions (e.g., social distancing).

This study is organized into five main sections. The paper starts with general information about the history and information of the disease. Section 2 provides a survey of the multiple forecasting techniques employed to predict the confirmed cases in the Indian context. Sections 3 and 4 discuss our findings and results. We conclude this research work in section 5 with the best performing forecasting model along with some interesting observations.

## 2 Technical Background

Forecasting techniques can be inculcated thereby assisting the government in designing better strategies and in making productive decisions. These techniques assess the situations of the past, enabling better predictions about the situation to occur in the future. Such predictions will help governments all over the world to prepare for the forthcoming situations. These types of forecasting techniques can play a very important role in yielding nearly accurate predictions.

We provide here a basic introduction to some of these techniques and concepts that were used in the research studies surveyed by us. For further details about different techniques, readers are advised to read research works pertaining to a particular used technique.

### 2.1 Regression Model

Regression analysis is a form of predictive modeling technique that investigates the relationship between dependent and independent variables. In statistics, linear regression is a linear approach to modeling the relationship between a scalar response (or dependent variable) and one or more explanatory variables (or independent variables).

Given a data set of *n* statistical units, a linear regression model [4] assumes that the relationship between the dependent variable *y* and the *p*-vector of regressors x is linear. This relationship is modeled through a disturbance term or error variable *ϵ* - an unobserved random variable that adds “noise” to the linear relationship between the dependent variable and regressors. Thus, the model takes the general form as shown by Eq. 1.

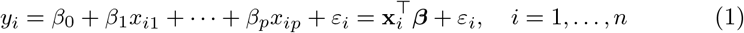

where *T* denotes the transpose, so that 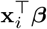 is the inner product between vectors *x_i_* and *β*.

Whereas, polynomial regression is a form of regression analysis in which the relationship between the independent variable *x* and the dependent variable *y* is modeled as an nth degree polynomial in *x*. Research work [9] used the polynomial regression for COVIE-19 predictions.

### 2.2 Auto-Regressive Integrated Moving Average (ARIMA) Model

ARIMA, short for ‘Auto-Regressive Integrated Moving Average,’ is a family of equations that describe a certain time series dependent upon their prior values, i.e. its own lags and lagged prediction errors, such that the equation can be used to estimate future values. The ARIMA process is also known as the Box-Jenkins method. The Box-Jenkins approach is for a merged ARIMA configuration to be fitted to a particular data set [10].

### 2.3 A Multilayer Perceptron

A multilayer perceptron (MLP) represents any type of feed-forward neural network composed of multiple hidden layers. MLPs [11] are being increasingly used in complex predicting tasks which can’t be solved using traditional machine learning methods. An MLP with a single hidden layer can be represented graphically as shown in Fig. 1. A one hidden-layered MLP can be represented in matrix notation by the following equation:

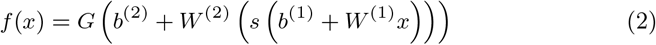

where *b*^(1)^, *b*^(2)^ are bias vectors, *W*^(1)^,*W*^(2)^ are weight matrices and *G* and *s* are activation functions. The vector 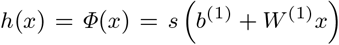 constitutes the hidden layer.

**Fig 1:**
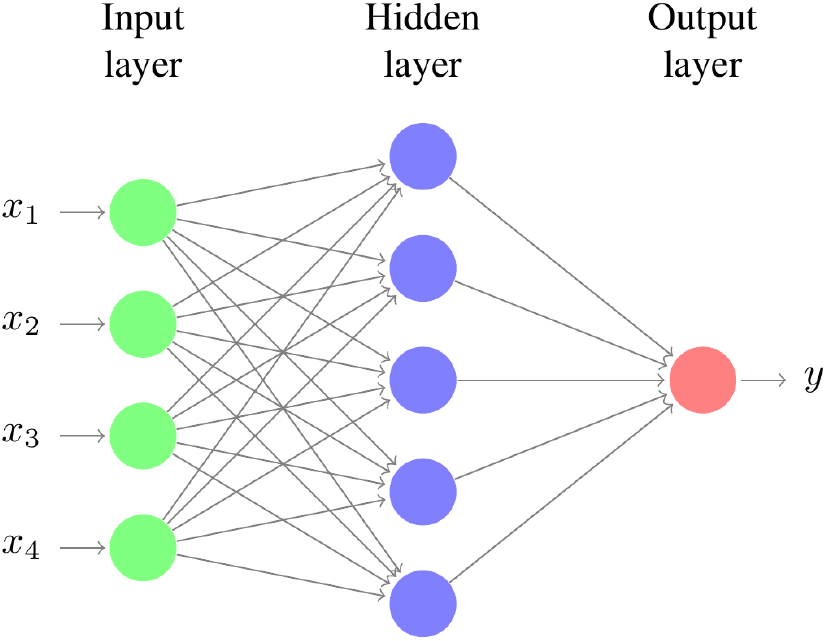
Multilayer perceptron with one hidden layer

### 2.4 Vector Autoregression

Vector Autoregression (VAR) [12] is a regression technique capturing the relationship of 2 time series which interact with each other. It is employed where the interdependence between the time dispositions included is bi-directional. The general equation for VAR model is:

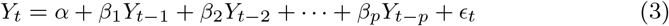

where *α* is the intercept, *β*_1_, *β*_2_,…*β_p_* are the coefficients of the lags of *Y* and *∊_t_* is the error considered as white noise.

### 2.5 Susceptible Exposed Infected Recovered (SEIR) Model

The SEIR model is constituted of four major factors which are - Susceptible (S) depicting the individuals which can get the disease, Exposed (E) referring to those people which have been already exposed to the disease, Infected (I) depicting the number of people which have been infected and can infect others and Recovered (R) which refers to those people who have became immune to the disease after contracting it once. Fig. 2 gives a pictorial representation of the SEIR Model [9].

**Fig 2:**
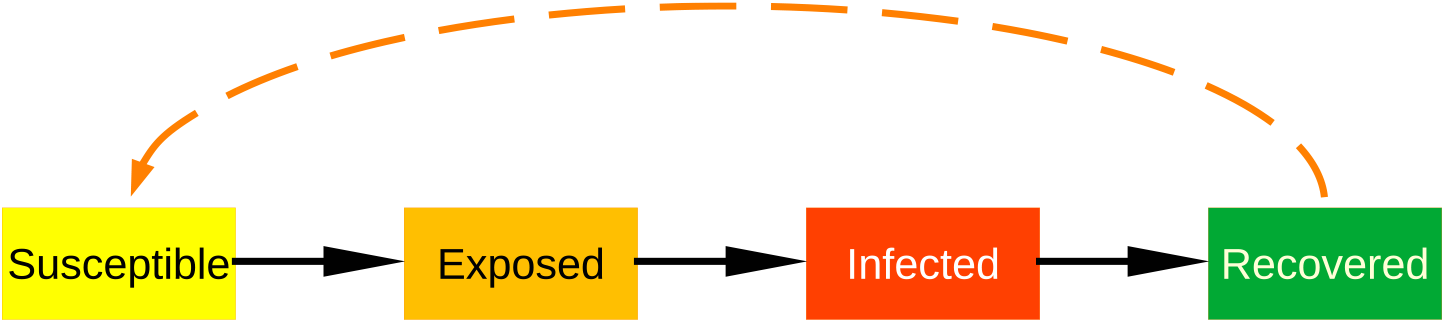
Graphical representation of SEIR model

### 2.6 Time Series Forecasting using Waikato Environment for Knowledge Analysis (Weka) Software

Time series analysis is the method of analyzing and describing a time-based sequence of data points using statistical techniques. Its forecasting involves the use of a formula to produce future event forecasts based on established events from the past. Time series data have a normal sequential order-different from traditional data mining and computer analysis applications where each data point offers an individual indication of the principle to be studied. Its topics include: capacity preparation, inventory refilling, demand projections and projected staffing ratios [13].

Weka was used to develop the forecasting model [13]. It is a series of data mining machine learning algorithms that can be implemented or named from the own Java code directly into a dataset. It provides resources to pre-process, identify, rectify, cluster, associate rules, and visualize results.

### 2.7 Recurrent Neural Networks (RNNs)

Recurrent neural networks (RNNs) fall in the domain of deep learning methods where the output from the previous step is fed as input to the current step. In traditional neural networks, its inputs and output data are independent of each other. But for tasks requiring the use of sequential information, we would require the information from previous computations done by the neural network [14].

Thus, RNNs came into existence, which possessed a memory like feature to remember the information from previously computed calculations. Theoretically, they can make use of this information in randomly long sequences but in practical cases they run into the vanishing gradient problem which inspired researchers to develop Long Short Term Memory Networks.

### 2.8 Long Short Term Memory Networks (LSTMs)

Long Short Term Memory (LSTM) was developed to deal with the vanishing gradient problem that can be encountered when training traditional RNNs. A common LSTM unit is composed of a cell, an input gate, an output gate and a forget gate. The cell remembers values over arbitrary time intervals and the three gates regulate the flow of information into and out of the cell. LSTM networks are well-suited to classifying, processing and making predictions based on time series data, since there can be lags of unknown duration between important events in a time series. The equations for the gates in a LSTM [14] are as follows:

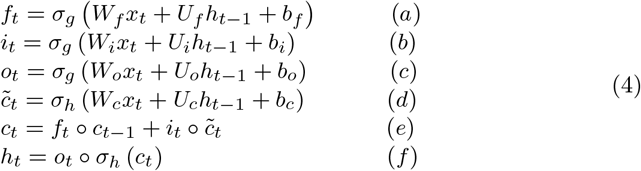

where *i_t_* represents input gate, *f_t_* represents forget gate, *o_t_* represents output gate, *h_t−_*_1_ is the output of the previous lstm block (at timestamp t-1), *x_t_* is the input at current timestamp, *b_x_* represents biases for the respective gates (x), *w_s_* refers to the weight for the respective gate (x) neurons and *σ* represents sigmoid function. Equation 4(a) is for the input gate, which tells us what new information we are going to store in the cell state (that we will see below). The second equation 4(b) is for the forget gate which tells the information to throw away from the cell state. The third one 4(c) is for the output gate which is used to provide the activation to the final output of the LSTM block at timestamp ‘t’. A block of LSTM at any point *t* is shown in Fig. 3.

**Fig 3:**
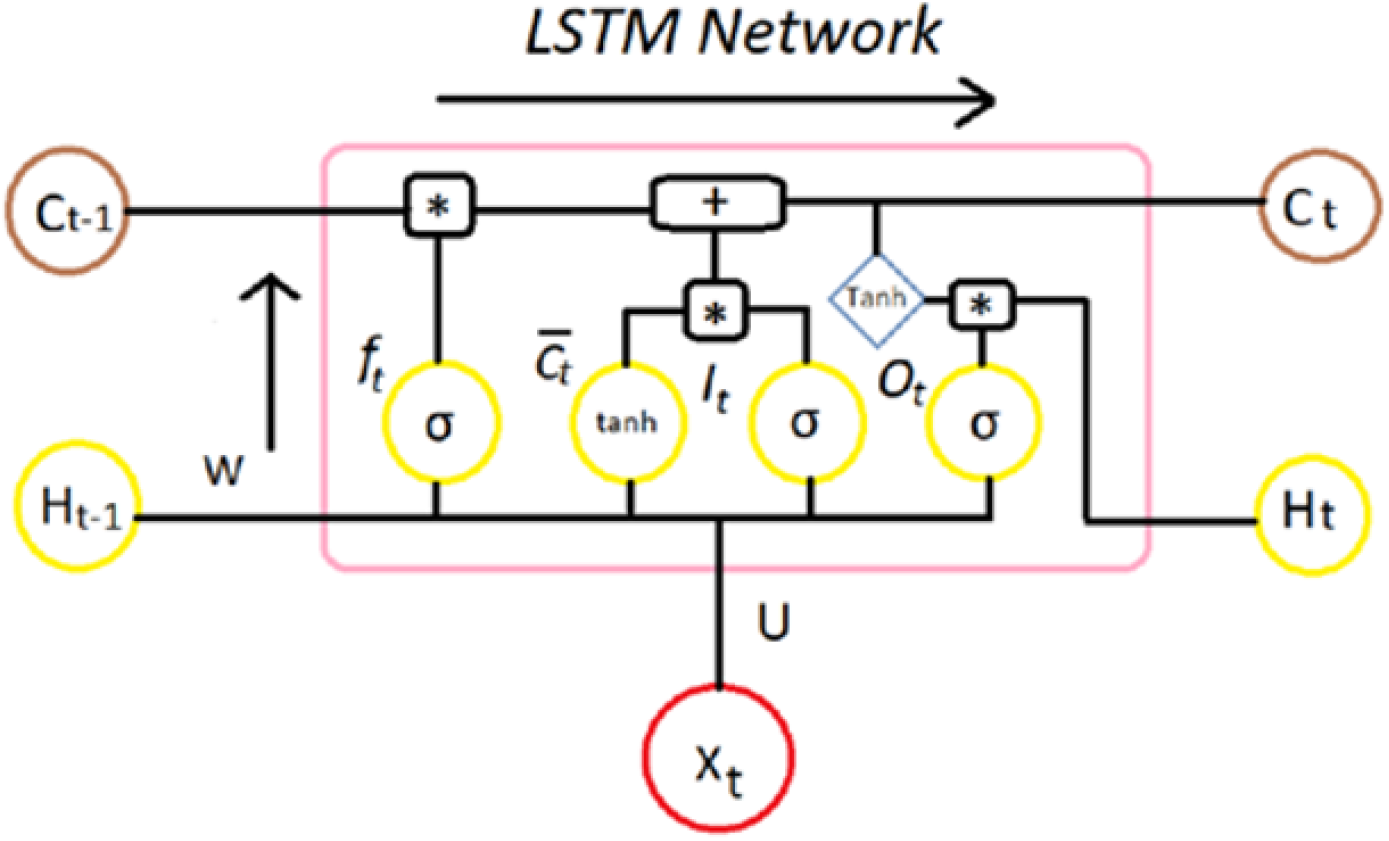
Block of LSTM network at some point of time ‘t’

### 2.9 Susceptible Infected Recovered (SIR) Model

SIR model is a disease modeling system like SEIR which segregates the whole community into 3 sections - Susceptible, Infected, and Recovered. The definitions of S, I, and R sections are the same as those of the SEIR model. The three categories are interrelated with each other and with parameters according to the following equations [15]: Rate of change of Susceptible Population is given by:

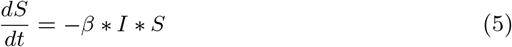

Rate of change of Infected Population is given by:

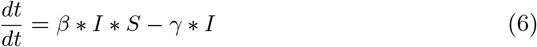

Rate of change of Recovered Population is given by:

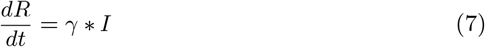

### 2.10 Curve Fitting

In this technique, we fit a curve or mathematical function that fits best to a sequence of input data values, conditioned upon some constraints. It can entail the use of interpolation, where an accurate fit to the data is a necessity. Otherwise, a smoothing function is adopted where only an approximate fit to the points is required [16]. Previous research [17] fitted the following polynomial function to their input data with least squares as their loss function:

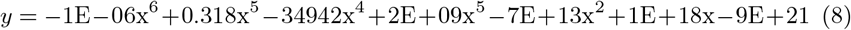

One [18] of the other researchers used an exponential fitting to predict the values. The equation employed is:

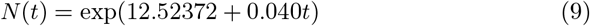

## 3 Analysis of Past Forecasting Models

### 3.1 Methodology Used

The analysis of each study is conducted by comparing their forecasted values for a certain duration with the actual (also called true) number of cases for that duration. This was implemented by extracting values from tables and graphs presented in these research works and then juxtaposing them with the values obtained from the Ministry of Health and Family Welfare website of Government of India [3]. We have used the Mean Absolute Percentage Error (MAPE) metric for contrasting between predicted and true values. MAPE was chosen because of its scale-independence property, which would remove the size of test data bias from the models. MAPE is a measure of how accurate a forecasting model is and gives its output as a percentage value. It is calculated by normalizing the average error at each point. This is done by dividing the errors by actual values and then averaging their sum using the total size of data using the following mathematical equation:

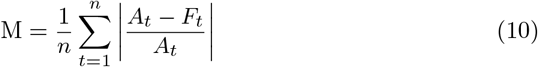

where *A_t_* is the actual value and *F_t_* is the forecast value.

### 3.2 Reviewed Past Research Works

For exhaustive research, we considered 10 research studies that focused on forecasting the number of COVID-19 confirmed cases in India. These research works were chosen to analyze a variety of forecasting techniques that are being used. These studies are shown in Table 1 along with some other information such as the models used by them, the source of their data, the duration of their forecasting, and the MAPE score they achieved.

**Table 1:** Reviewed Research Works

| S. No. | References | Forecasting Techniques | Duration of Forecasting | Data Source | MAPE score |
| --- | --- | --- | --- | --- | --- |
| 1 | [9] | SEIR and Regression models | 20 days | JHU [23] | SEIR: 25.533, Regression: 21.889 |
| 2 | [10] | ARIMA and NAR | 50 days | [3] and [30] | ARIMA: 362.1761 |
| 3 | [11] | LR, MLP and VAR | 69 days | Kaggle [22] | LR: 1745454.432, MLP: 80.057, VAR: 43289.290 |
| 4 | [13] | Time Series Forecasting using Weka | 24 days | Kaggle [25] | 55.489 |
| 5 | [14] | LSTM | 25 days | Govt. Of India [26] | 63.357 |
| 6 | [19] | Genetic programming-based model (GP) [GEP model] | 10 days | Mendley Time series datasets [29] | 7.827 |
| 7 | [20] | Exponential Growth Model and ML based LR model | 33 days | Humanitarian website for COVID data | LR: 662.441, Exponential: 2096.000 |
| 8 | [18] | Short term forecasting using exponential curve fitting | 15 days | [27] | 6.4807 |
| 9 | [15] | SIR Model | 36 days | WHO [28] | 66.819 |
| 10 | [17] | Least square fitted model | 35 days | [27] | 39.816 |

## 4 Results

In this section we demonstrate how well these models fared against the true number of cases in India. This information is shown by 2 different plots for each study. First plot represents both the predicted and true curve against the predicted number of days while the second plot represents the relationship between the forecasted and true data. Ideally, second plot should tend to achieve linearity.

Fig. 4 represents the results of Linear Regression technique proposed by the researchers of [11]. This research used the official number of cases from January 22, 2020 to April 10, 2020 as their input data and predicted values for a duration of 69 days starting from April 11, 2020 to June 18, 2020. This model scores really low on our metric with a MAPE score of 1745454.432. The error for some points reaches upto the magnitude of 10^11^. It means that this research was very far off from the true value of confirmed cases that actually took place.

**Fig 4:**
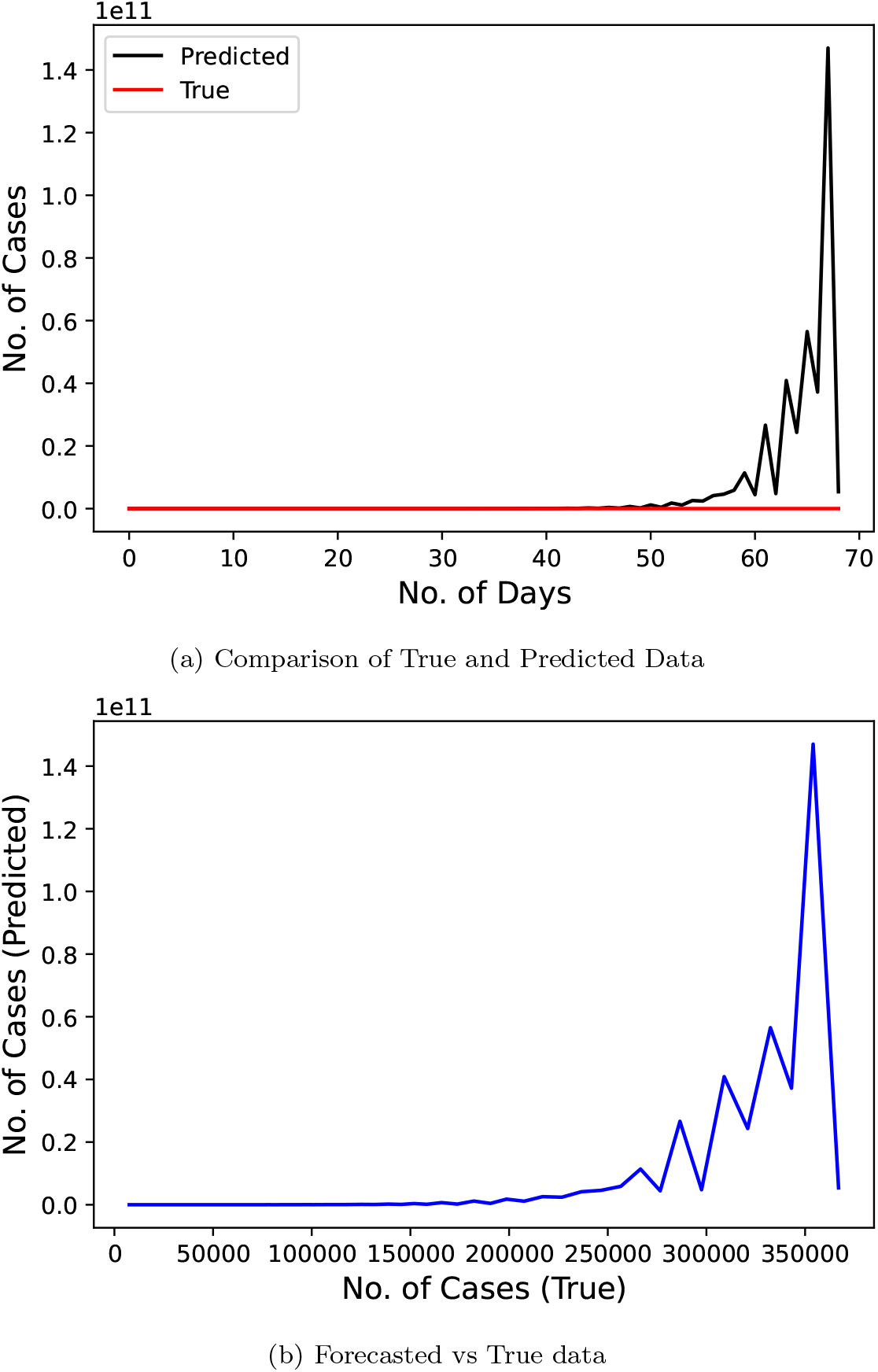
Confirmed COVID-19 cases comparison derived from research [11]

Fig. 5 represents the results of the ARIMA model used by the researchers [10]. This research predicted the number of cases for a duration of 50 days starting from March 5, 2020 to April 23, 2020 using the data from January 31, 2020 to March 4, 2020. This model performs moderately with a MAPE score of 66.819.

**Fig 5:**
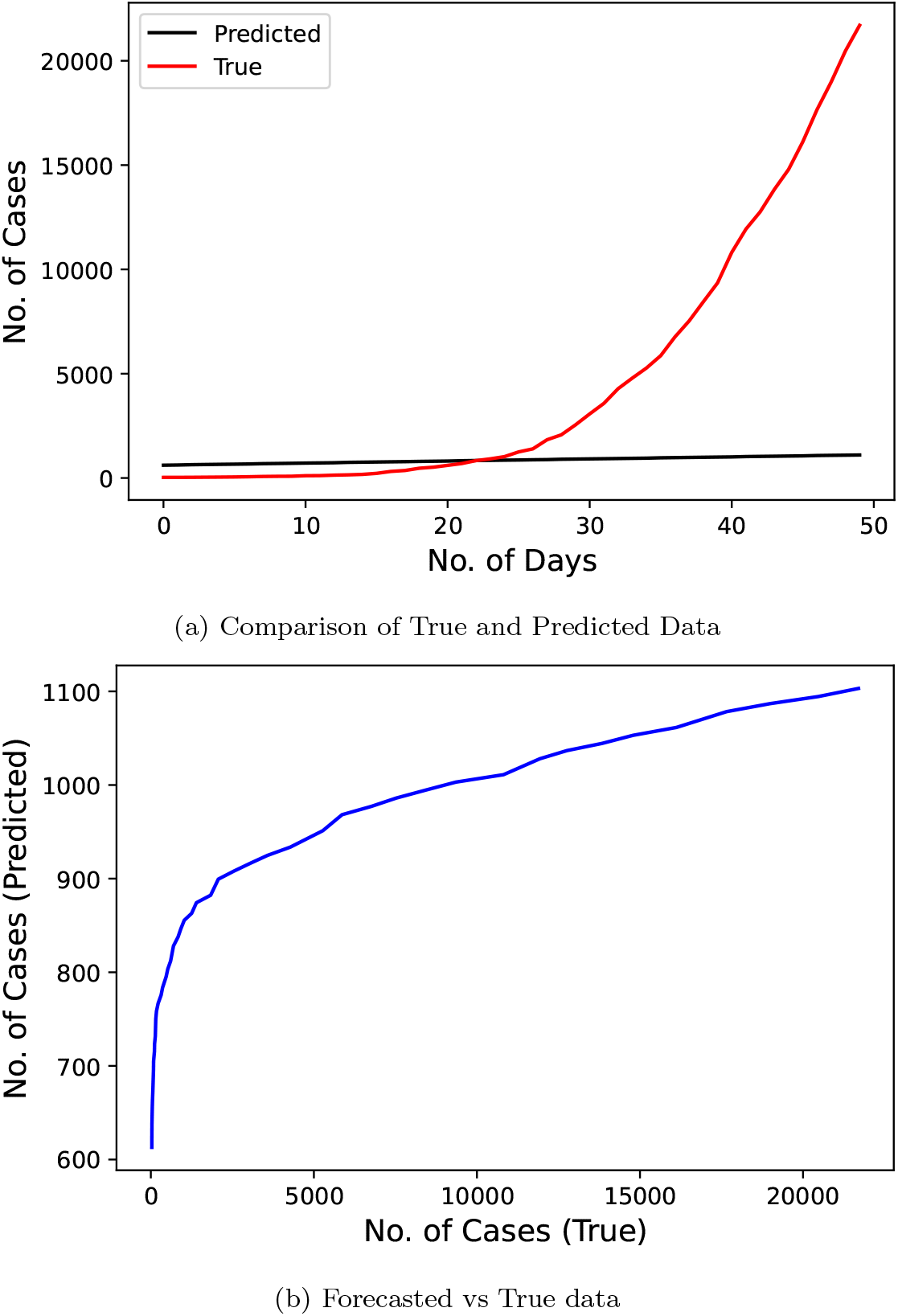
Confirmed COVID-19 cases comparison derived from research [10]

Fig. 6 represents the results of Multilayer Perceptron model proposed by the researchers [11]. Duration of both the training and testing data are same as that of the Linear Regression model. This model performed much better than the LR technique with a MAPE of 80.057.

**Fig 6:**
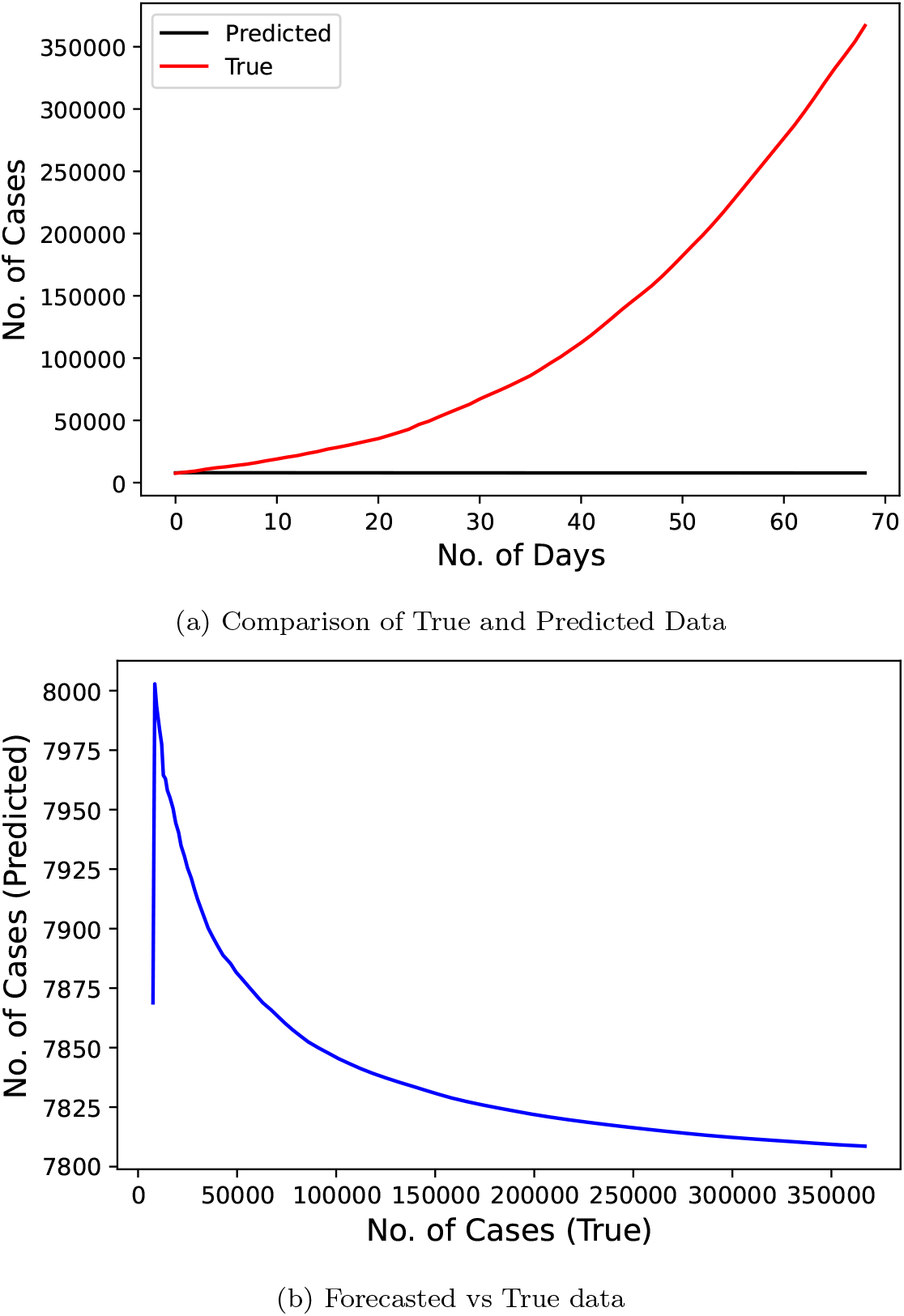
Confirmed COVID-19 cases comparison derived from research [11]

Fig. 7 shows the results of Vector Autoregression technique proposed by the researchers [12] on the same input data as previous two techniques. This model again gives a poor performance with MAPE of 43289.29.

**Fig 7:**
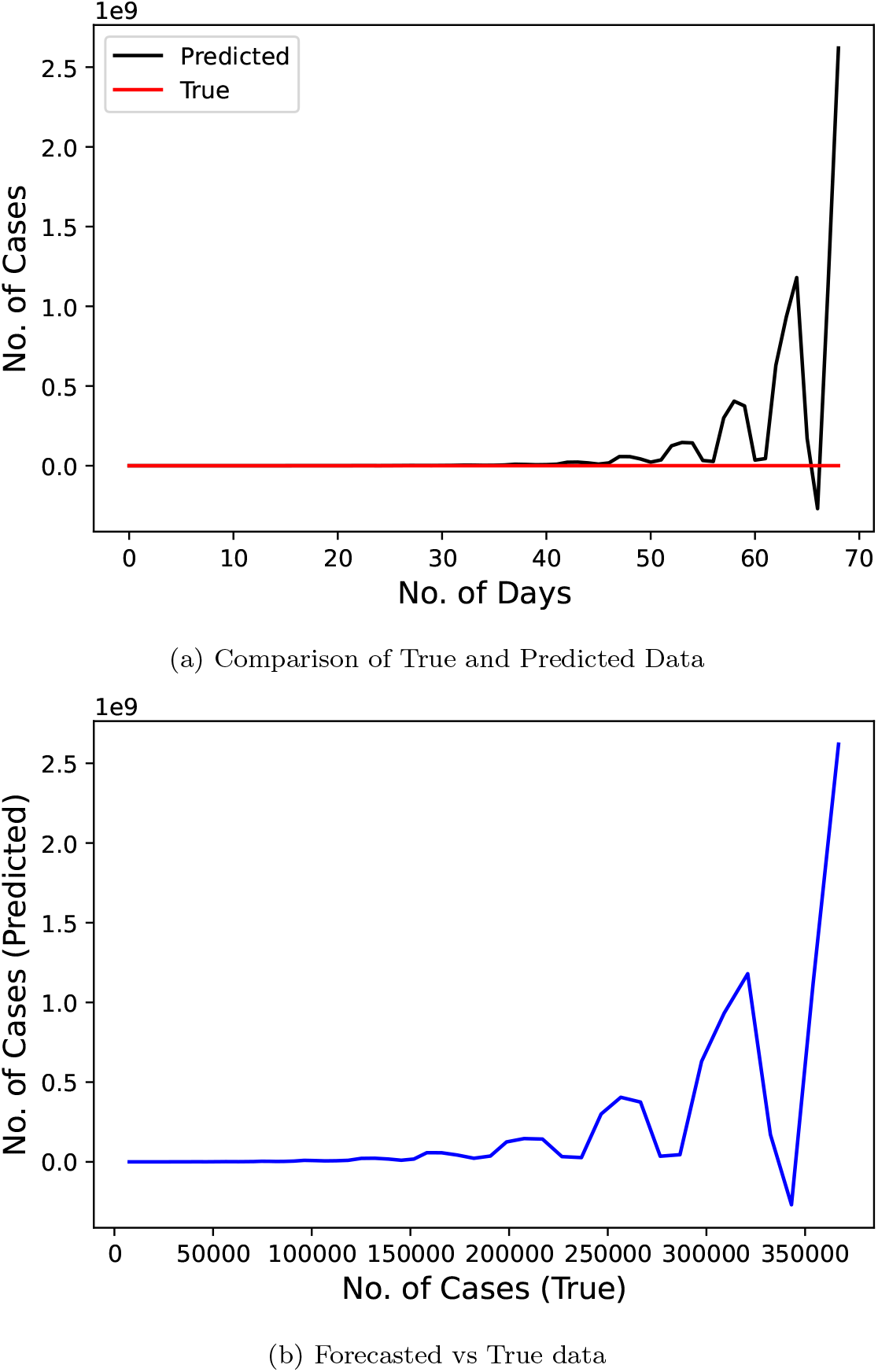
Confirmed COVID-19 cases comparison derived from research [12]

Fig. 8 displays the results of time series forecasting conducted by the researchers [13]. Researchers used the official number of cases from January 22, 2020 to April 3, 2020 to train their model. The predicted duration was of 24 days starting from April 4, 2020 to April 27, 2020. This study scores a MAPE score of 55.48984343.

**Fig 8:**
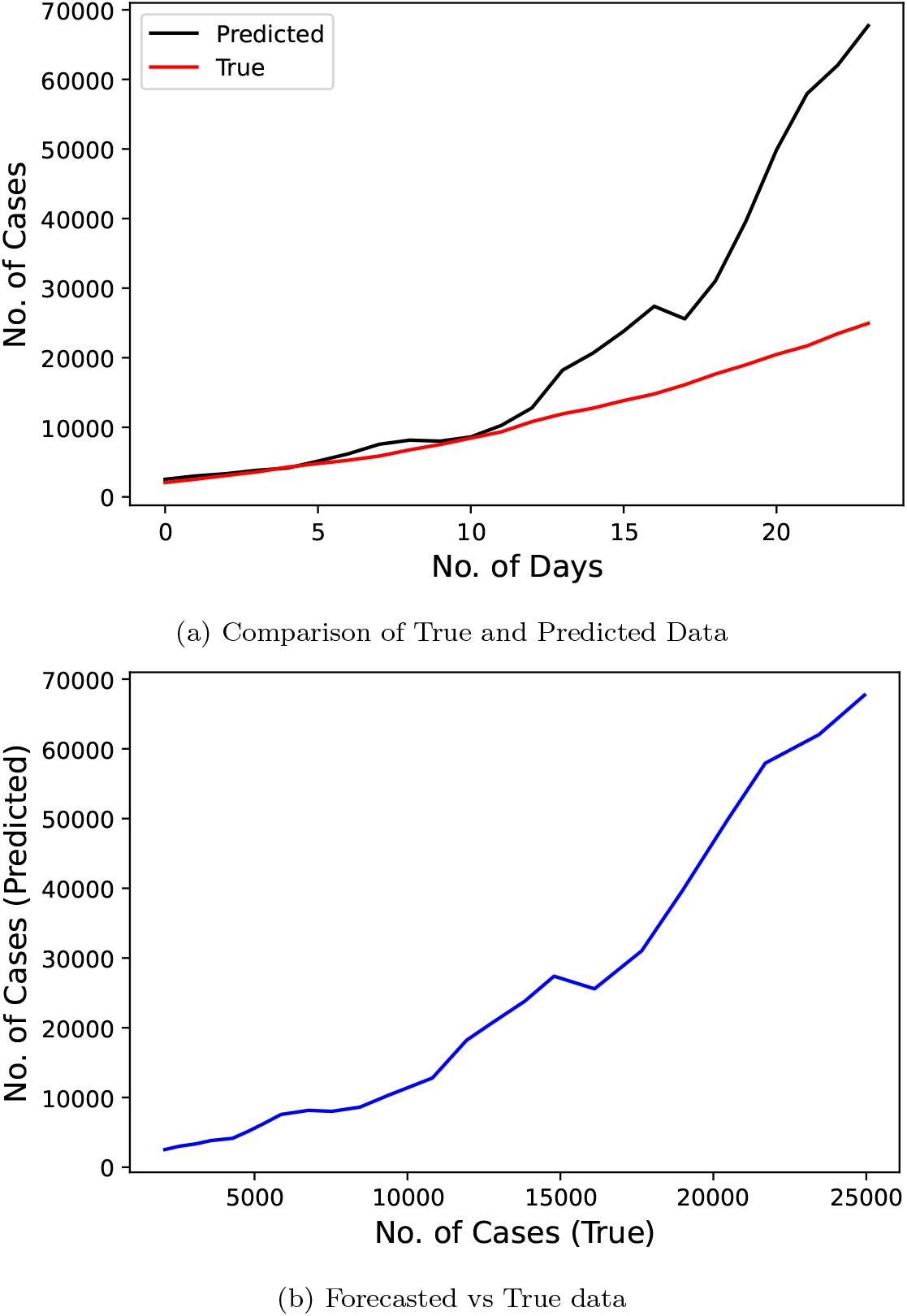
Confirmed COVID-19 cases comparison derived from research [13]

Fig. 9 represents the results of the LSTM model used by the researchers [14] to predict for a duration of 25 days from April 5, 2020 to April 29, 2020. The researchers took the data starting from January 30, 2020 to April 4, 2020 for their model. This study scored good on MAPE metric with a value of 63.357.

**Fig 9:**
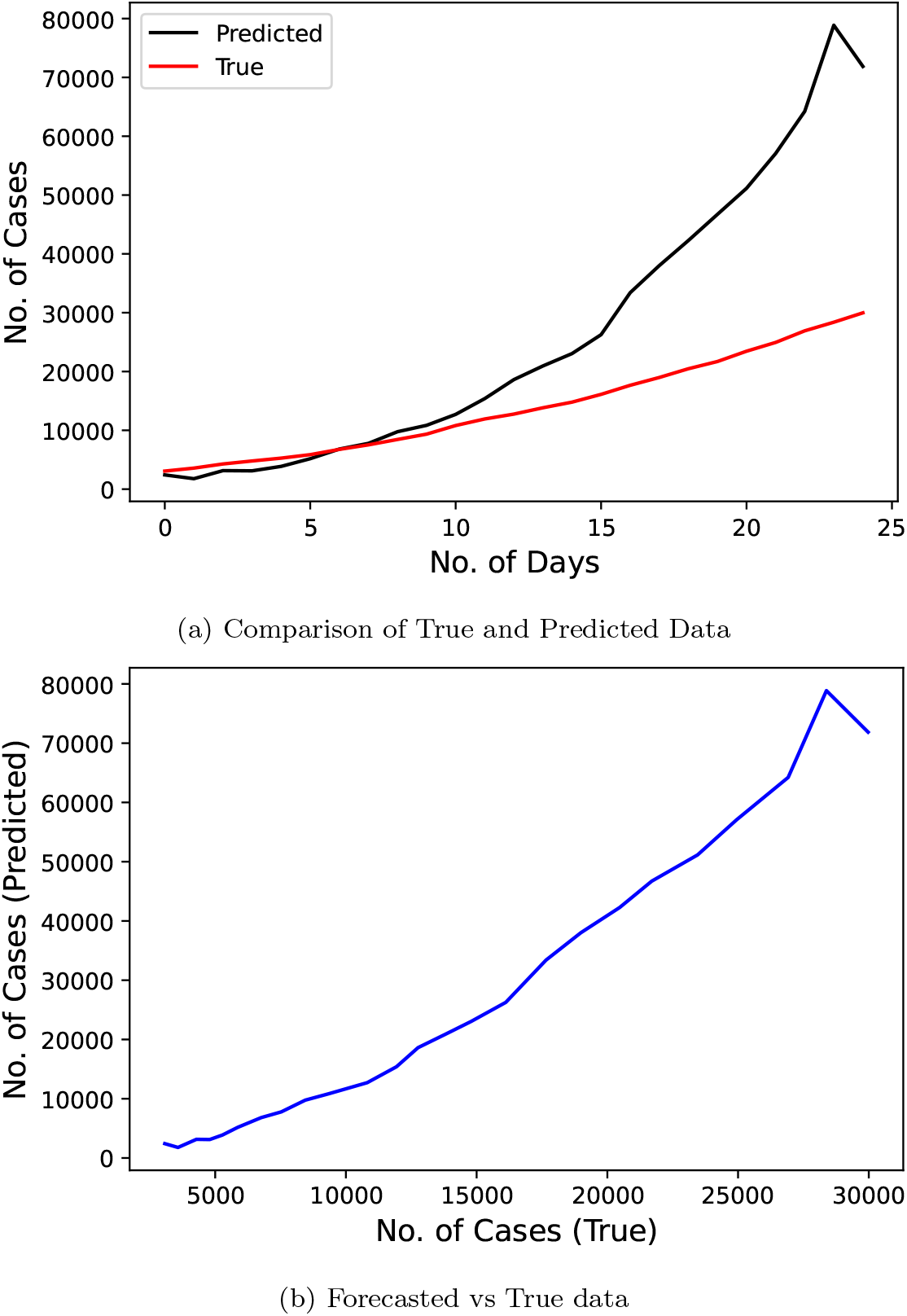
Confirmed COVID-19 cases comparison derived from research [14]

Fig. 10 represents the results of GEP model proposed by the researchers [19]. The said research predicted the number of infected cases from May 14, 2020 to May 23, 2020 for a duration of 10 days. Input for that research model were the number of official cases from March 24, 2020 to May 13, 2020. This model got a very good MAPE score of 7.827.

**Fig 10:**
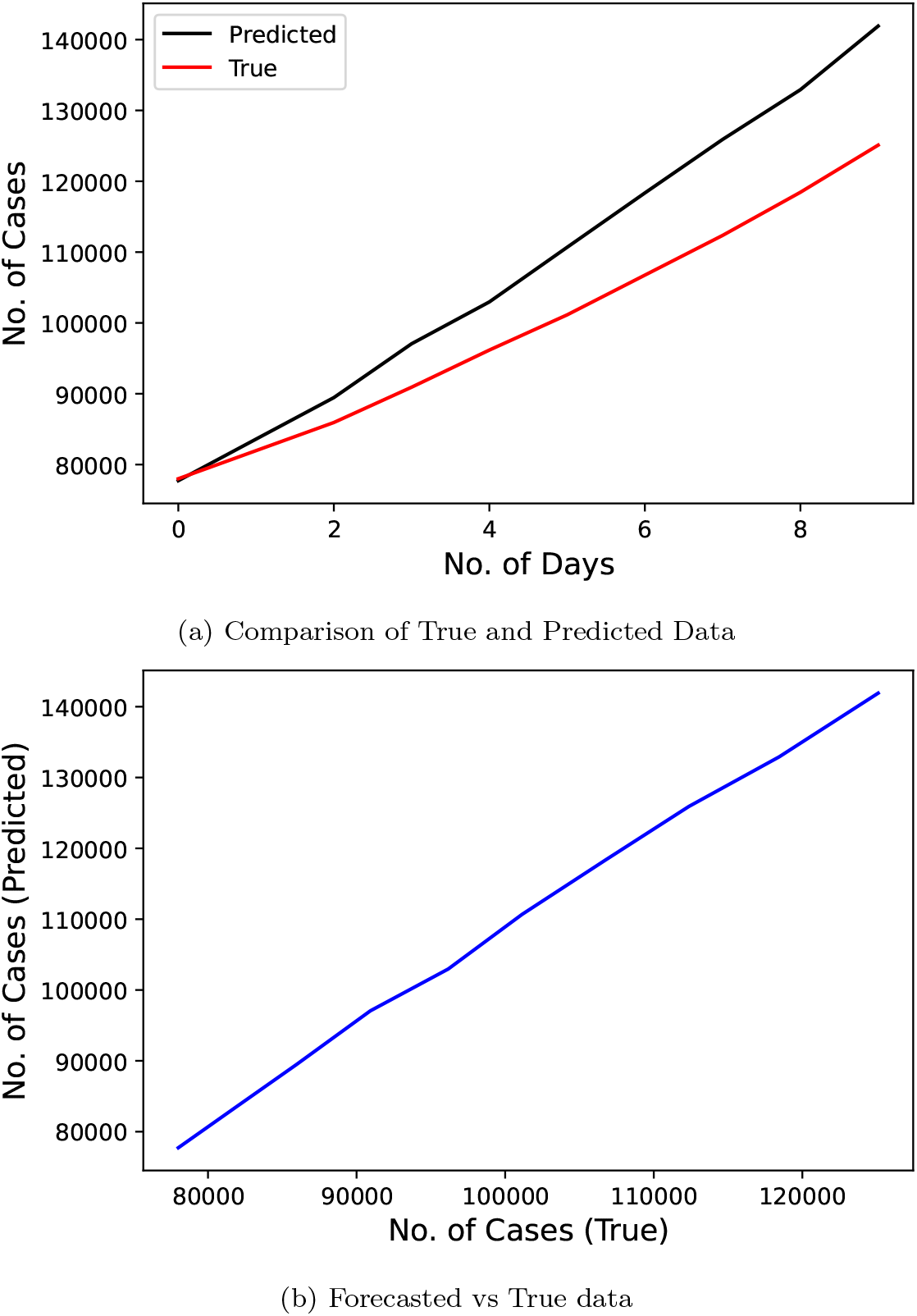
Confirmed COVID-19 cases comparison derived from research [19]

Fig. 11 demonstrates the results of Linear Regression technique proposed by the researchers [20] and the research used the official number of cases from February 11, 2020 to May 11, 2020 as the input data to train the model. The predicted duration was of 33 days starting from May 12, 2020 to June 30, 2020. This model scores quite low on our metric with a score of 662.441.

**Fig 11:**
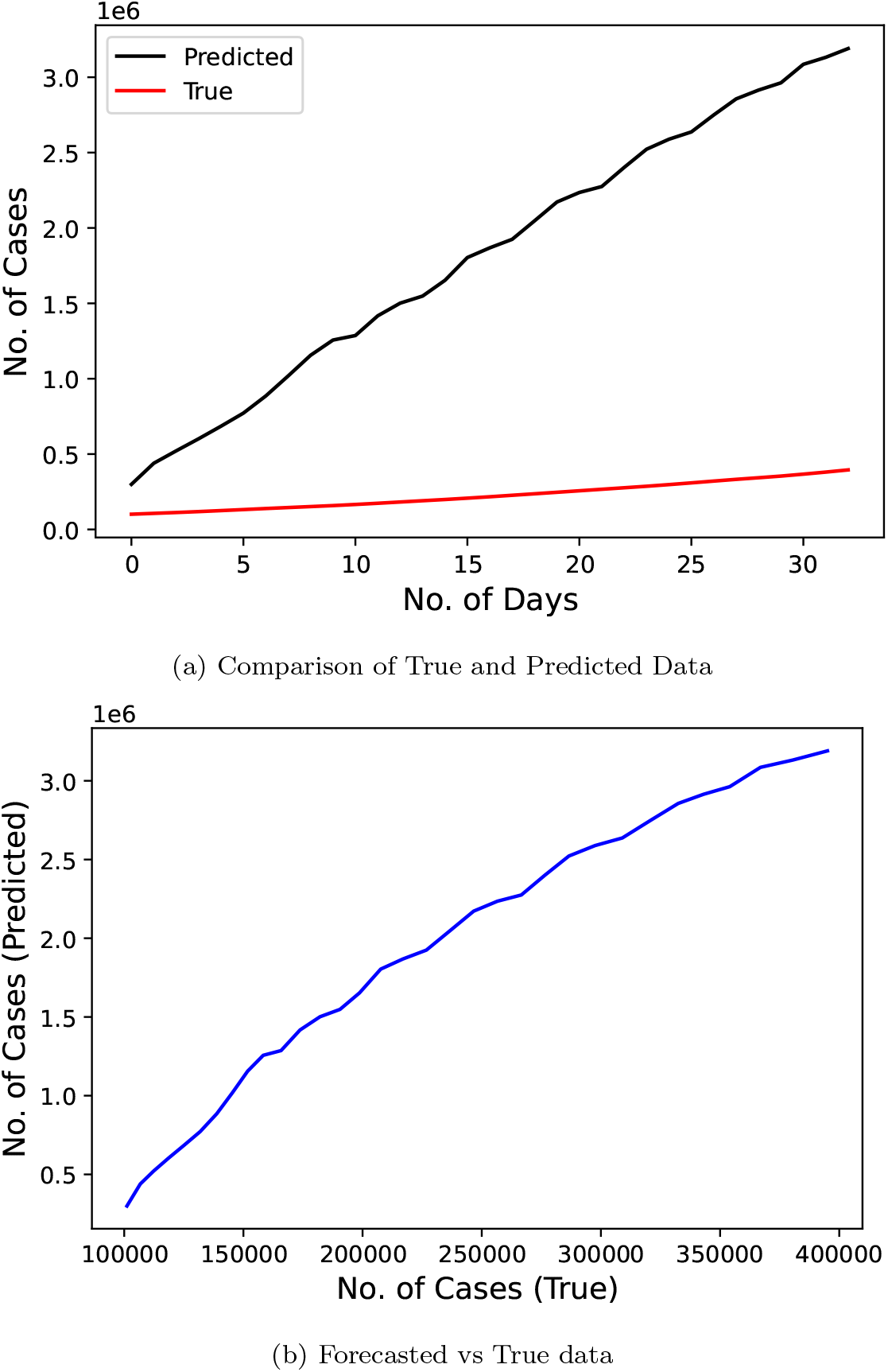
Confirmed COVID-19 cases comparison derived from research (LR) [20]

Fig. 12 displays the results of the exponential model used by the researchers [20]. Size of both training and test data remains analogous to the linear regression technique. This model gives MAPE metric result of 2096.000.

**Fig 12:**
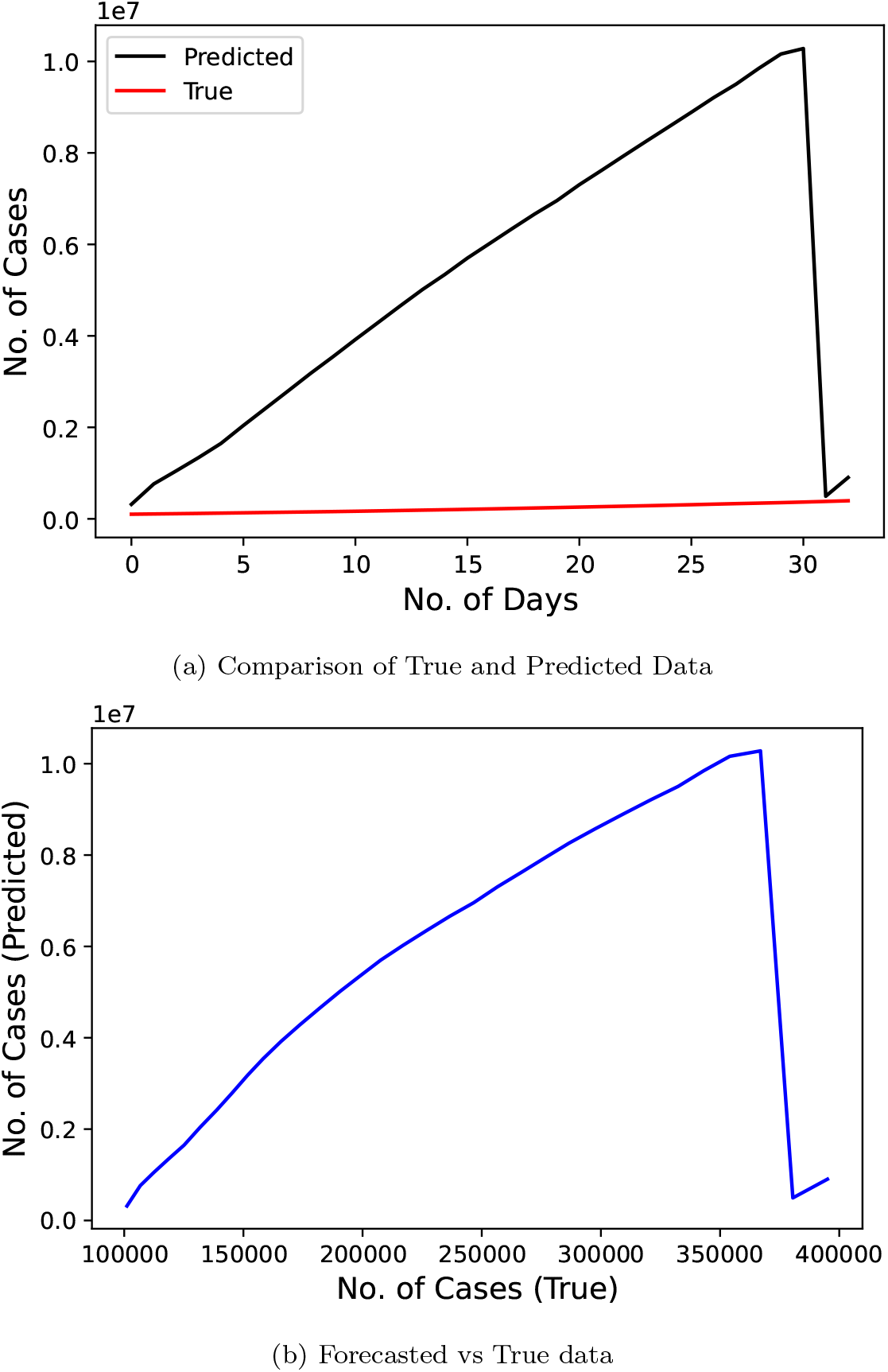
Confirmed COVID-19 cases comparison derived from research (exponential model) [20]

Fig. 13 demonstrates the results of SEIR model proposed by the researchers [9] who used the official number of cases from January 30, 2020 to March 30, 2020 as input data to train the model. The predicted duration was of 20 days staring from March 31, 2020 to April 13, 2020. This model scores good on the MAPE metric with a score of 25.533.

**Fig 13:**
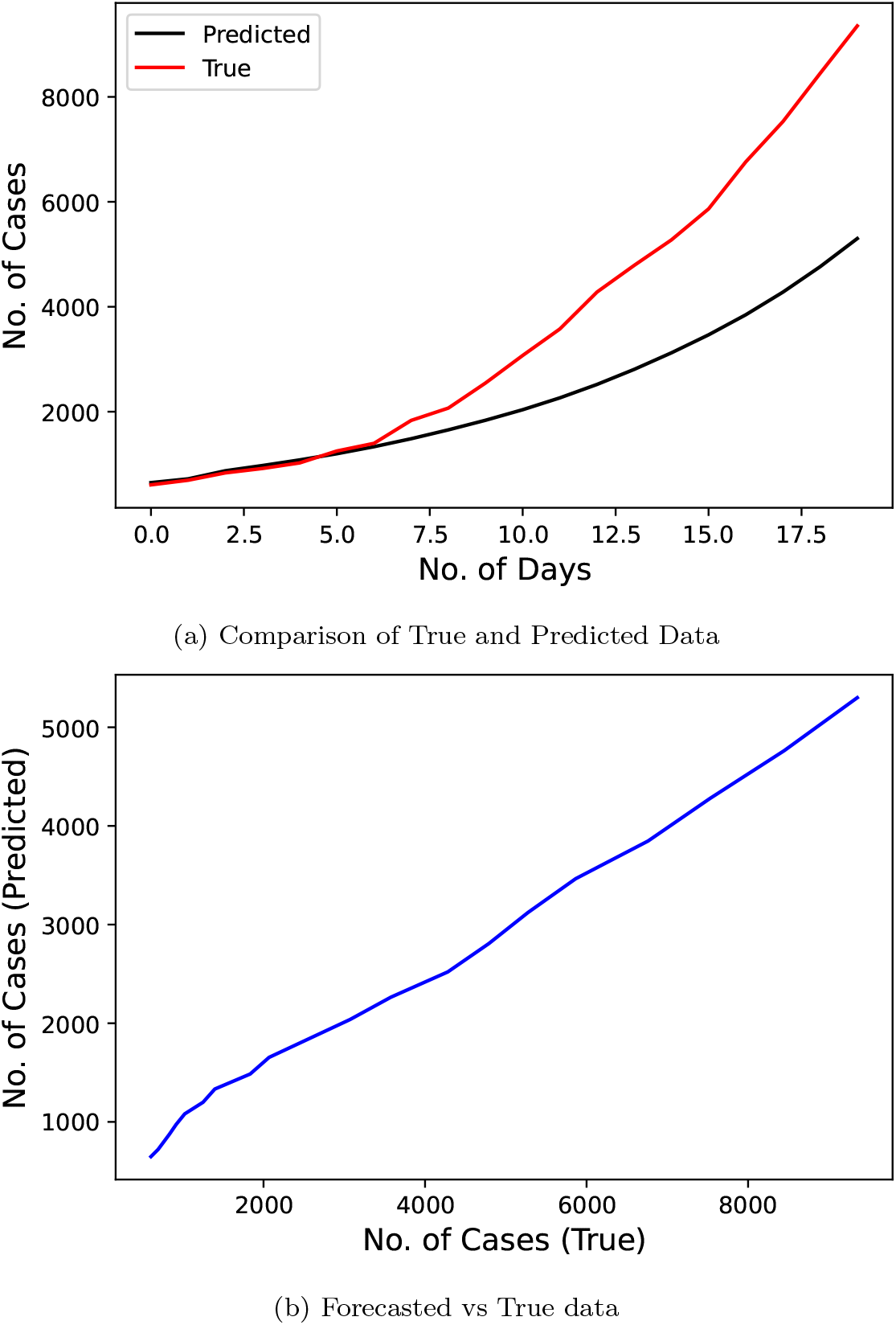
Confirmed COVID-19 cases comparison derived from research (SEIR model) [9]

Fig. 14 represents the results of Regression model proposed by the researchers [19]. Duration of both the training and testing data are same as that of the SEIR model. Also, this model performed better than the SEIR technique with a MAPE of 21.889.

**Fig 14:**
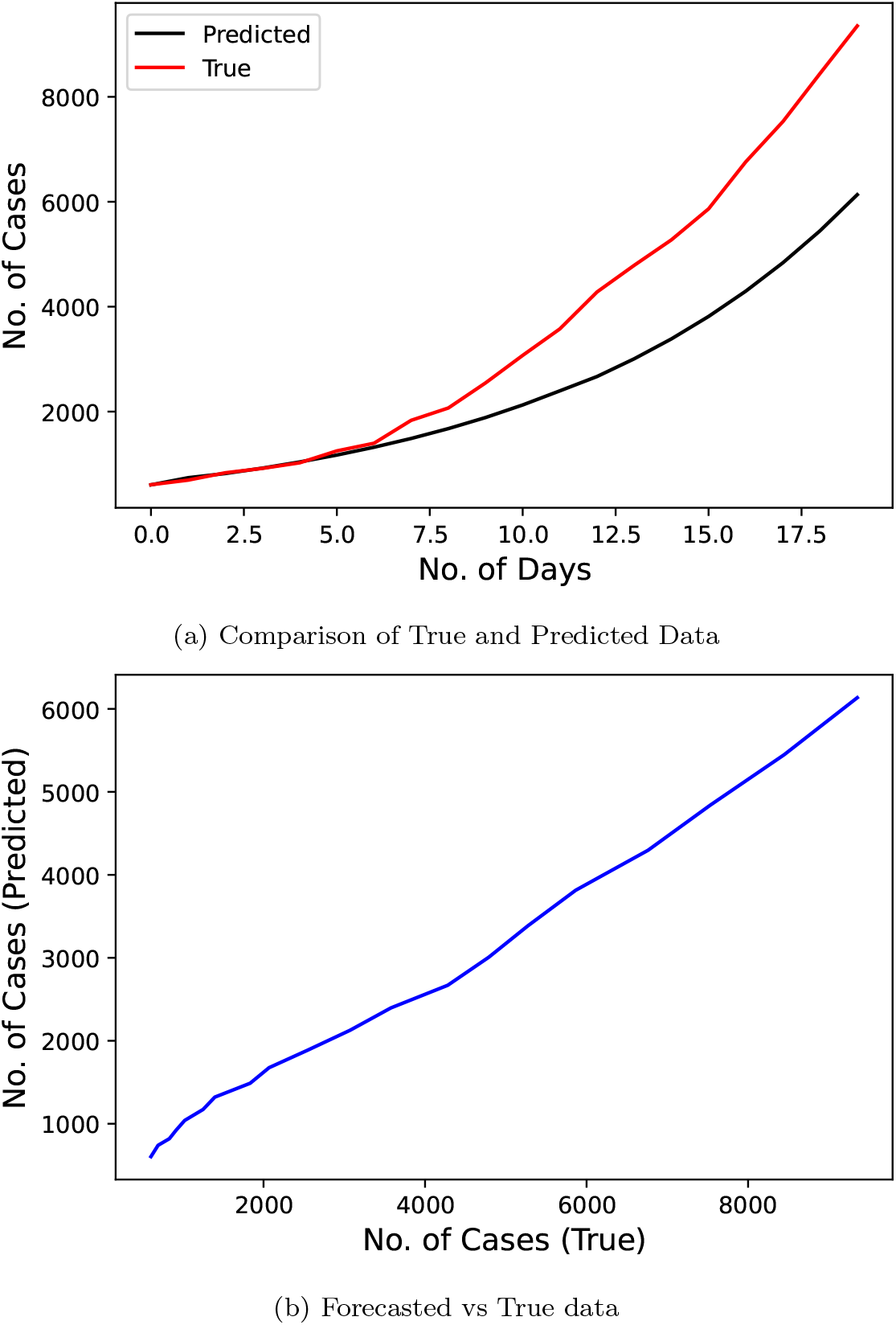
Confirmed COVID-19 cases comparison derived from research (regression model) [19]

Figure 15 represents the results of Least Square Fitted model proposed by the researchers [17] and this research used the official number of cases from January 30, 2020 to April 26, 2020 as the input data to train the model. The predicted duration was of 35 days starting from April 27, 2020 to May 31, 2020. This model gives a MAPE score of 39.816.

**Fig 15:**
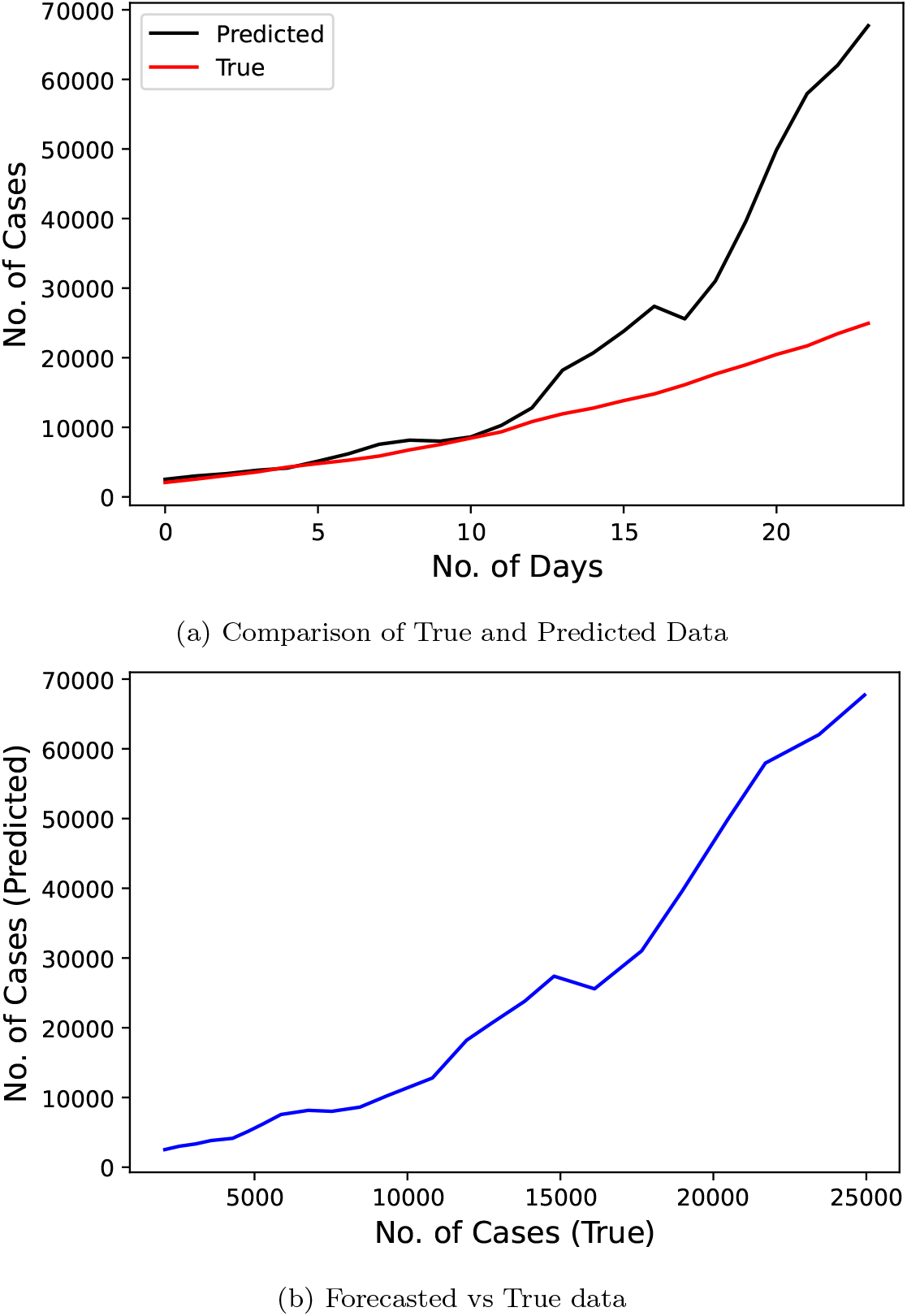
Confirmed COVID-19 cases comparison derived from research [17]

Fig. 16 represents the results of the SIR model used by the researchers [15] and predicted the number of cases for a duration of 36 days starting from March 23, 2020 to April 27, 2020 using the data from March 3, 2020 to March 22, 2020. This model scores a modicum MAPE score of 66.819.

**Fig 16:**
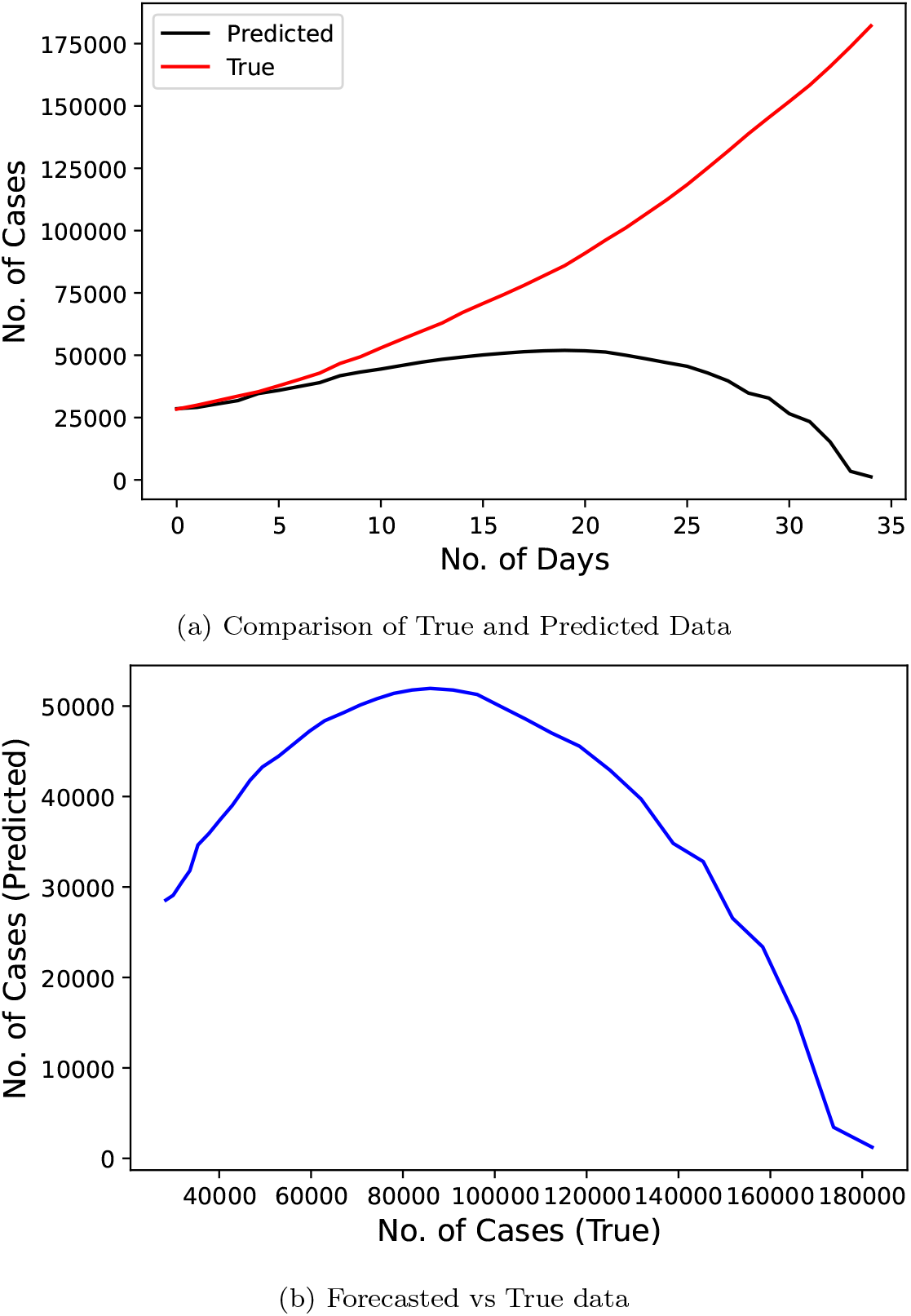
Confirmed COVID-19 cases comparison derived from research (SIR model)

Figure 17 represents the results of Linear Regression technique proposed by the researchers [18] who used used the official number of cases from June 1, 2020 to June 10, 2020 as the input data to train the model. The predicted duration was of 15 days starting from June 10, 2020 to June 24, 2020. This model scores best on the MAPE metric with a score of 6.4807.

**Fig 17:**
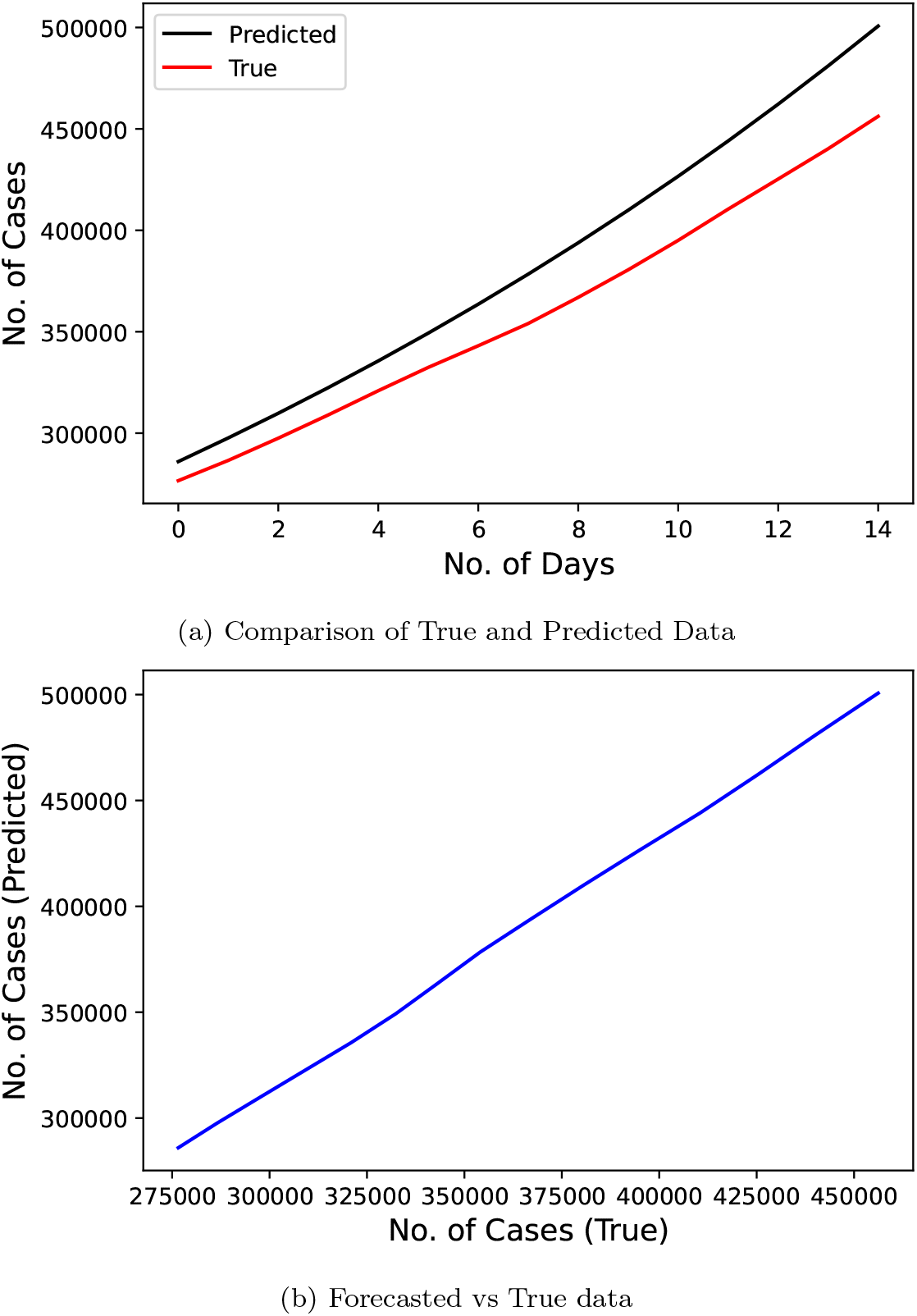
Confirmed COVID-19 cases comparison derived from research (LR model) [18]

## 5 Conclusion

This research work considered data related to daily confirmed cases of COVID-19 in India during lockdowns on national level. From the results shown in the previous section, we observe that even after using a size independent metric, models which predicted for lesser durations (less than 20 days) [19], [18] scored best on our MAPE metric. Overall, exponential curve fitting [18] on recent data produced the lowest value score of 6.4807. But its use should be limited to only short-term forecasting as predicting values for a longer duration will lead to the fitted curve moving away from the true values.

Discarding these outlier models which conducted forecasting for smaller durations, we conclude that the least square fitted model [17] outperforms all the other techniques and thus, should be used for future purposes of predicting the infection rate under similar conditions. Also, disease modeling systems such as SEIR and SIR [9], [15] fared much better than other regression models, and we believe that if applied with more precision, they can achieve good results.

While we compared the prediction performance among selected models, we could not recommend one particular model over another, for two reasons. First, in most cases, the models were mostly far from accurate. Second, in addition to the algorithm used for prediction, assumptions of the algorithms and baseline conditions should also be considered. This is particulary important when such risk prediction is to be used to revise health policies and recommend practices for the public, such as lockdown, safety distance, and reopening. National lockdowns, for example, are often associated with substantial economic and societal costs. According to the Interional Monetary Fund [6], many countries have spent roughly 20 percent of their GDP to support emergency responses during the COVID-19 pandemic. As the daily infection rate of the coronavirus continues to increase in many countries such as India, Brazil, and the U.S., it is likely due to a combination of factors, such as delayed public response and inaccurate projections. Findings from this study can inform future development of risk prediction models of infectious disease, particularly by considering both virus-intrinsic characteristics and external variables, such as mass testing, social distancing, and decision on lockdown or reopening of institutions.

## Data Availability

Data is already available in the paper's references.

## References

1. https://covid19.who.int accessed on July 31, 2020

2. https://www.who.int/emergencies/diseases/novel-coronavirus-2019/question-and-answers-hub/q-a-detail/q-a-coronaviruses accessed on July 31, 2020

3. Ministry of Health and Family Welfare, https://www.mohfw.gov.in accessed on July 31, 2020

4. Montgomery, Douglas C., Elizabeth A. Peck, and G. Geoffrey Vining. “Introduction to linear regression analysis,” Vol. 821. John Wiley & Sons, 2012.

5. https://www.washingtonpost.com/politics/2020/08/05/health-202-why-individual-models-coronavirus-deaths-are-often-wrong/ accessed on August 8, 2020

6. https://www.bbc.com/news/business-52450958 accessed August 8, 2020

7. https://www.pnas.org/content/117/28/16092 accessed August 8, 2020

8. https://www.bbc.com/news/business-51706225 accessed August 8, 2020

9. Gaurav Pandey, Poonam Chaudhary, Rajan Gupta, Saibal Pal, “SEIR and Regression Model based COVID-19 outbreak predictions in India”, doi: 10.1101/2020.04.01.20049825

10. Farhan Mohammad Khan, Rajiv Gupta, “ARIMA and NAR based prediction model for time series analysis of COVID-19 cases in India,” Journal of Safety Science and Resilience, Volume 1, Issue 1, Pages 12–18, 2020

11. R. Sujath, Jyotir Moy Chatterjee & Aboul Ella Hassanien, “A machine learning forecasting model for COVID–19 pandemic in India” Stochastic Environmental Research and Risk Assessment, volume 34, pages 959–972(2020), doi: 10.1007/s00477-020-01827-8

12. https://www.machinelearningplus.com/time-series/vector-autoregression-examples-python/

13. Sunita Tiwari, Sushil Kumar, Kalpna Guleria, “Outbreak trends of CoronaVirus (COVID19) in India: A Prediction” Disaster Med Public Health Prep. 2020 Apr 22: 1–6., doi: 10.1017/dmp.2020.115

14. Anuradha Tomar, Neeraj Gupta. “Prediction for the spread of COVID-19 in India and effectiveness of preventive measures” Science of The Total Environment Volume 728, 1 August 2020, 138762, doi: 10.1016/j.scitotenv.2020.138762

15. Jay Naresh Dhanwant, V. Ramanathan, “Forecasting COVID 19 growth in India using Susceptible-Infected-Recovered (S.I.R) model”, arXiv:2004.00696 [q-bio.PE]

16. https://en.wikipedia.org/wiki/Curve_fitting

17. Mr. Sudip Ghosh, “An Overview: Situation Assessment and Prediction of Corona Virus in India” Mukt Shabd Journal Volume IX Issue V, MAY/2020 Issn No: 2347–3150

18. Hemanta Kumar Baruah, “Nearly Perfect Forecasting of the Total COVID-19 Cases in India: A Numerical Approach”, doi: 10.1101/2020.06.13.20130096

19. Rohit Salgotra, Mostafa Gandomi, Amir H Gandomi, “Time Series Analysis and Forecast of the COVID-19 Pandemic in India using Genetic Programming” Chaos, Solitons & Fractals Volume 138, September 2020, 109945, doi: 10.1016/j.chaos.2020.109945

20. Ajit Kumar Pasayat, Satya Narayan Pati, Aashirbad Maharana, “Predicting the COVID19 positive cases in India with concern to Lockdown by using Mathematical and Machine Learning based Models” doi: 10.1101/2020.05.16.20104133

21. Shinde, G.R., Kalamkar, A.B., Mahalle, P.N. et al. Forecasting Models for Coronavirus Disease (COVID-19): A Survey of the State-of-the-Art. SN COMPUT. SCI. 1, 197 (2020). https://doi.org/10.1007/s42979-020-00209-9

22. https://www.kaggle.com/imdevskp/corona-virus-report/data

23. https://coronavirus.jhu.edu/

24. WHO. Situation report–1. Novel coronavirus (2019-nCoV). https://www.who.int/emergencies/diseases/novel-coronavirus-2019/situation-reports

25. Kaggle. Novel corona virus 2019 dataset, https://www.kaggle.com/sudalairajkumar/novel-corona-virus-2019-dataset

26. https://www.mygov.in/covid-19/?cbps=1

27. https://www.worldometers.info/

28. https://www.who.int

29. https://data.mendeley.com/datasets/tmrs92j7pv/1

30. http://covid19india.org/

